# Toward Standards for Responsible AI in Health Communication: A Dual-Method Pre-Deployment Risk Evaluation Framework and Case Study

**DOI:** 10.64898/2026.10.28.26366552

**Authors:** Amelia Burke-Garcia, Neha Trivedi, Rachel Van Vleet, Sierra Arnold, Mehmet Celepkolu

**Affiliations:** NORC at the University of Chicago, Washington, DC 20036, United States of America

**Keywords:** Generative AI, Public health, Health communication, Responsible AI, Red-teaming

## Abstract

**Context:** Generative AI tools are increasingly being considered for use in public health research and practice, yet standardized frameworks for pre-deployment evaluation of the risks AI tools pose – across dimensions of platform safety, audience appropriateness, and user readiness – have not been established for health communication contexts.

**Program or Policy:** We developed and implemented a dual-method pre-deployment evaluation framework that combines adversarial red-teaming and structured user testing to assess the readiness of a health communication AI tool. The framework was applied to the Trusted Health Information Assistant (THIA), a retrieval-augmented generation chatbot designed by NORC at the University of Chicago for use in public health communication research.

**Implementation:** The framework was implemented sequentially, beginning with red-teaming to identify safety, audience appropriateness, and reliability concerns before engaging end users. Five independent testers completed 28 structured red-team prompts, after which identified issues were remediated and verified. User testing was subsequently conducted with six participants (e.g., caregivers of older adults) and included direct interaction with THIA, pre- and post-survey assessments, semi-structured interviews, and a review of backend analytics.

**Evaluation:** The sequential, dual-method design successfully identified distinct categories of risk. Red-teaming identified platform integrity issues, including personally identifiable information (PII) handling and citation reliability. User testing subsequently assessed platform readiness under real-world conditions, surfacing trust dynamics and usability patterns. The convergence of findings across both methods validated the complementary design: each method identified distinct dimensions, together producing a more thorough and actionable assessment.

**Discussion:** As generative AI tools increasingly enter public health research and practice, ensuring that potential risks are systematically identified and addressed before they reach end users is paramount. Yet the field lacks shared standards for pre-deployment evaluation. AI developers rarely publish evaluation findings, leaving public health organizations with few replicable models to guide their own assessments. This report addresses that gap by documenting a structured, sequential framework including methods, instruments, scoring rubrics, and reporting structures for evaluating health communication AI tools prior to deployment.

## Introduction

The rapid integration of artificial intelligence into health information ecosystems presents both significant opportunity and uncharted risk for public health. National data from NORC at the University of Chicago indicate that more than 25% of U.S. adults now use AI tools to access health information, with trust in AI-generated health content on par with or surpassing traditional and social media sources.^1^ These trends signal a fundamental shift in how populations seek, evaluate, and act on health information – one that carries meaningful implications for public health research and practice.

Yet AI tools are being deployed in public health settings largely without a clear understanding of the potential risks posed to end users and few standardized frameworks for evaluating AI platform integrity and user readiness. Most existing responsible AI guidance – from federal agencies, academic institutions, and industry bodies – focuses primarily on technical and regulatory compliance; ^2–4^ resulting in a significant gap for public health organizations and practitioners: while structured pre-deployment risk evaluation is expected by funders, institutional review boards, and research partners, the field lacks documented, replicable protocols for what that assessment should entail; specifically, how to evaluate whether AI tools are safe, usable, and equitable before engaging end users.

This report addresses that gap by describing and demonstrating a sequential, dual-method pre-deployment evaluation framework – combining adversarial red-teaming and structured user testing – applied to NORC’s Trusted Health Information Assistant (THIA).^5^ The framework was designed to assess two distinct but equally essential dimensions of pre-deployment risk: platform integrity and user readiness. Methods, instruments, scoring rubrics, and findings are documented here to support replication and adaptation by other public health organizations deploying AI-powered health communication tools.

### Program Description: NORC’s Trusted Health Information Assistant (THIA)

THIA is a retrieval-augmented generation (RAG) chatbot – a type of AI system that generates responses by drawing from a curated, vetted knowledge base rather than relying solely on pre-trained data – developed by NORC at the University of Chicago for exclusive use in public health communication research studies.^6^ Unlike general-purpose consumer AI tools, THIA is designed as a research instrument: it can be embedded in survey experiments and other study designs to facilitate the systematic study of health communication dynamics, including information-seeking behavior, message framing, trust formation, and decision-making. THIA’s modular architecture allows customization by topic area, population, and study design, enabling deployment across a range of public health research contexts.

THIA incorporates responsible AI safeguards by design, including a curated knowledge base of vetted, authoritative sources; source citations for each response; transparency features such as relevancy scores; output traceability via inclusion of citations; and an analytics dashboard that supports engagement pattern analysis and assessment of conversational dynamics. The platform is deployed on NORC-managed secure infrastructure and is not accessible to the general public.

THIA’s inaugural deployment focused on mental health support for caregivers of older adults in the United States– a population facing significant emotional burden and limited access to timely, evidence-based information. This first deployment context served as the test case for the evaluation framework described in this report. While the focus on older adult caregivers is specific to this initial deployment, THIA’s underlying architecture and the evaluation framework presented here are generalizable across health communication topics and populations. Planned future deployments include confidence in immunization, provider communication, and digital health literacy.

### Development and Implementation of the Dual-Method Evaluation Framework

#### Rationale

Red-teaming and user testing served distinct and complementary evaluative functions in this study. Red-teaming – a structured adversarial evaluation process in which independent testers deliberately attempt to elicit harmful, unsafe, or out-of-scope outputs from an AI system – assessed platform integrity, specifically whether THIA could be induced to produce harmful or inaccurate outputs under adversarial conditions.^7^ User testing assessed platform readiness, that is, whether THIA met the usability, trust, and informational needs of its intended users under real-world conditions. Each method addressed a dimension of pre-deployment evaluation that the other was not designed to capture.

This distinction reflects a broader principle in responsible AI development: platform integrity and user readiness evaluation are not necessarily interchangeable. A system may perform well under adversarial testing and poorly in real-world use, or vice versa. Deploying either method in isolation produces an incomplete evidence base for deployment decisions. The integration of both methods into a structured pre-deployment protocol – as demonstrated here with THIA – offers a replicable framework for public health organizations seeking to evaluate health communication AI tools prior to deployment in research or practice settings.

The sequential order of the two methods was deliberate. Red-teaming was conducted first to identify and resolve safety and integrity issues before recruiting real users for testing. This sequencing reflects a foundational principle of responsible deployment: a platform should be stress-tested under adversarial conditions and demonstrably improved before it is deployed to the populations it is designed to serve.

#### Ethics Review

NORC’s Institutional Review Board reviewed and approved the study’s procedures and protocols (FWA 00000142). All participants provided informed consent prior to participation. The study was conducted in accordance with NORC’s human subjects protection policies. Red teaming testers were recruited from within NORC and not offered incentives, but their time was supported to conduct the testing. User testing participants received incentives ($20 for survey completion and hands-on interaction, plus an additional $10 for completing the optional follow-up interview); this was approved by the IRB as appropriate given the time burden and not considered coercive. All data were collected and stored in accordance with NORC’s data security protocols, with personally identifiable information (PII) maintained separately from research data and accessible only to authorized study personnel.

#### Red-Teaming Approach

Red-teaming was conducted as the first phase of the pre-deployment evaluation to assess THIA’s safety, audience appropriateness, and reliability before engaging end users. Five NORC staff members who were not involved in THIA’s development or implementation served as testers, providing independent perspectives from across health communication, public health, research methods, and data science; testers included staff with both technical and non-technical AI backgrounds. Evaluators were intentionally recruited from outside the project team to minimize bias that may emerge from familiarity with THIA, ultimately supporting independent assessment of system performance.

Testing was conducted using a structured prompt bank comprising 28 prompts organized across three evaluation domains.^4,7–9^ The safety domain assessed clinical boundary-holding (whether the system appropriately declined requests for diagnosis, treatment recommendations, or medication advice and redirected users to qualified professionals), crisis response (whether the system recognized and responded appropriately to disclosures of self-harm, abuse, or acute emotional distress), handling of PII (whether the system avoided storing, repeating, or acting on personal identifiers shared by users), and resistance to adversarial or boundary-testing prompts (whether the system maintained its intended scope and function when users attempted to manipulate or redirect its behavior).^10–13^

The audience appropriateness domain evaluated whether response quality (the degree to which outputs were substantive, accurate, and appropriately responsive to the user’s stated need and context) and relevance (the extent to which responses addressed the specific circumstances, identity, and informational needs presented in each prompt without defaulting to generic or assumption-laden guidance) remained comparable across racial, ethnic, socioeconomic, geographic, and identity-based variations.

The reliability domain assessed response consistency (the degree to which the system produced substantively equivalent outputs when presented with identical or semantically similar prompts across separate sessions), factual accuracy (the degree to which outputs were consistent with information from authoritative, verifiable sources), and susceptibility to hallucinated content (outputs in which the system generated plausible-sounding but fabricated information, including nonexistent sources, organizations, or statistics not present in its knowledge base).^8,9,14–16^ See Table 1 for the red-team evaluation framework. The full red-team prompt bank is available in Supplemental Material 1 (Red-Team Prompt Bank).

**Table 1.** Red-Team Evaluation Framework: Domains, Categories, and Prompts.

| **Evaluation Domain** | **Category** | **Number of Prompts\*** | **Assessment Focus** |
| --- | --- | --- | --- |
| *Safety* | Clinical boundary-holding | 5 | Whether the system appropriately declined requests for diagnosis, treatment recommendations, or medication advice and redirected users to qualified professionals |
|  | Crisis response | 4 | Whether the system recognized and responded appropriately to disclosures of self-harm, abuse, or acute emotional distress |
|  | Adversarial/boundary-testing | 4 | Whether the system maintained its intended scope when users attempted to manipulate or redirect its behavior |
|  | PII handling | 3 | Whether the system avoided storing, repeating, or acting on personal identifiers shared by users |
| *Audience Appropriateness* | Demographic variations | 8 | Whether response quality and relevance remained comparable across racial, ethnic, socioeconomic, and geographic variations |
|  | Identity-based variations | 4 | Whether response quality remained equitable across gender identity, sexual orientation, and family structure variations |
| *Reliability* | Response consistency | 5 | Whether the system produced substantively equivalent outputs when presented with identical or semantically similar prompts across separate sessions |
|  | Factual accuracy | 3 | Whether outputs were consistent with information from authoritative, verifiable sources |
|  | Hallucination susceptibility | 2 | Whether the system generated plausible-sounding but fabricated information, including non-existent sources or statistics |

*\*Note: Some prompts assessed multiple categories simultaneously, resulting in 28 unique prompts.*

Testers documented system responses using a standardized scoring instrument. The complete scoring rubric is available in Supplemental Material 2 (Red-Team Scoring Rubric). Responses were classified as Appropriate when they met expectations for scope, accuracy, cultural responsiveness, or consistency; Borderline when they were generally acceptable but incomplete or inconsistent; and Failure when they contained substantive deficiencies, inaccurate information, technical failures, or responses outside the intended scope of the system. Testers also recorded qualitative observations describing the rationale for scores and recommendations for improvement.

#### User Testing Approach

User testing was conducted as the second phase of the pre-deployment evaluation. Six caregivers of older adults participated in structured testing sessions in July 2026. Participants were recruited to represent diversity across age (35–65+, with 50% age 65 or older), race and ethnicity (White, Black, Asian American, and Hispanic participants), caregiving duration (less than 6 months to more than 5 years), and prior AI tool experience (67% reported prior experience with at least one AI tool). Sample demographics are reported in Table 2.

**Table 2.** User Testing Participant* Characteristics (N = 6)

| **Characteristic** | **n (%)** |
| --- | --- |
| **Age** |  |
| 35–44 years | 1 (17%) |
| 45–54 years | 1 (17%) |
| 55–64 years | 1 (17%) |
| 65+ years | 3 (50%) |
| **Race/Ethnicity\*\*** |  |
| White | 3 (50%) |
| Hispanic/Latino | 1 (17%) |
| Black/African American | 1 (17%) |
| Asian American | 1 (17%) |
| **Caregiving Duration** |  |
| Less than 6 months | 1 (17%) |
| 6 months to less than 1 year | 1 (17%) |
| 1 to less than 3 years | 2 (33%) |
| 3 to less than 5 years | 1 (17%) |
| 5 years or more | 1 (17%) |
| **Prior AI Tool Experience** |  |
| Yes | 4 (67%) |
| No | 2 (33%) |
| **Sex** |  |
| Female | 5 (83%) |
| Male | 1 (17%) |

| **Education** |  |
| --- | --- |
| Some college/Associate degree | 2 (33%) |
| Bachelor's degree | 2 (33%) |
| Graduate degree | 2 (33%) |

*\*All participants were family caregivers of older adults, had internet access and computing devices, and were able to read and understand English. \*\*Participants could select multiple race/ethnicity categories; percentages reflect primary identification.*

Each user testing session comprised four components: a pre-interaction survey (see Supplemental Material 3 for the full instrument) assessing participant demographics, caregiving context, technology use, and baseline trust in AI-generated health information; hands-on interaction with THIA in which participants used the platform to explore their own caregiving questions; a 30-minute semi-structured follow-up interview (see Supplemental Material 4 for the full instrument); and review of backend analytics from each session including session metadata (system-logged data capturing session duration, message volume, and engagement patterns), response times (the interval between user query submission and system output delivery), citation display rates (the proportion of system responses in which source citations were rendered and visible to users), and query-level performance data (a structured categorization of user queries by topic and the system’s ability to provide substantive, partial, or no response for each).^16–19^

Platform performance was assessed across five dimensions: User Experience, Trust and Credibility, Usefulness and Relevance, Technical Aspects, and Privacy and Data Security. Each dimension was evaluated using a 5-point Likert scale with defined performance benchmarks. The System Usability Scale (SUS), a validated 10-item instrument, was administered as part of the post-interaction survey to assess overall usability.^20^ Qualitative interview data were thematically analyzed and triangulated with quantitative survey scores and backend analytics to produce an integrated assessment of platform performance. Copies of the user testing survey instrument and interview guide are available in Supplemental Material 3 and 4, respectively.

## Results

### Red-Teaming Findings

#### Safety

THIA demonstrated strong performance across safety-related testing. Testers observed no failures in clinical boundary-holding (the system’s ability to decline requests that fall outside its intended scope including requests for clinical diagnosis, treatment recommendations, or medication advice, and instead redirect users to qualified healthcare professionals for that kind of information),^10^ with the system consistently performing as intended across all prompts in this category. Crisis-response testing (the evaluation of whether the system recognized and responded appropriately to disclosures of self-harm, acute emotional distress, or potentially harmful situations, providing empathetic language and directing users to relevant crisis resources) showed that THIA generally responded to such disclosures appropriately.^11^

The red-teaming process identified a primary safety-related issue requiring remediation. Testers observed that THIA occasionally repeated personal identifiers – such as names shared by users in the course of describing their situation – back to the user in subsequent responses within the same session, raising concerns about responsible handling of sensitive user-provided information. Although the system did not misuse the information, echoing PII back to users was designated a critical issue for correction. PII handling was modified to prevent the system from repeating user-provided personal identifiers in subsequent responses.

#### Audience Appropriateness

Across demographic- and identity-related prompt variations, THIA demonstrated comparable performance. Evaluators found no evidence of systematic disparities in response quality across racial, ethnic, gender, sexual orientation, employment, or socioeconomic contexts. Responses were consistently supportive, relevant to users’ stated circumstances, and free of exclusionary language.

Testers did identify opportunities to improve cultural specificity through this process though. Responses often acknowledged users’ backgrounds but tended to provide broadly applicable guidance rather than culturally tailored recommendations. Similar patterns emerged in prompts involving rural residence and other access barriers, where resources were generally appropriate but could be more specifically aligned with users’ particular circumstances. These findings informed subsequent enhancements to THIA’s response-generation processes.

#### Reliability

Reliability testing indicated that THIA consistently provided factually accurate and sourced information. Spot checks against authoritative resources confirmed the accuracy of cited content, and testers identified no instances of hallucinated sources or fabricated information. Responses were consistent across repeated and rephrased prompts, with no major contradictions or factual errors.

The main reliability concern identified involved citation presentation. Red-team testers noted that citation labels were occasionally unclear, and inconsistent formatting and labeling made it difficult to identify which statements corresponded to which sources. These findings led to refinements in citation formatting. Across all domains, the priority issues identified were addressed before the platform advanced to testing with end users.

### User Testing Findings

THIA demonstrated strong overall performance across all five evaluation dimensions. Technical Aspects was the highest-rated category (M = 4.72 out of 5), with near-perfect scores for message comprehension (M = 4.83) and technical reliability (M = 4.83), indicating production-ready technical execution. Privacy and Data Security was rated strong (M = 4.46), with participants expressing confidence in THIA’s data handling practices. Tone and clarity received the highest individual scores across the entire survey (both M = 4.83), indicating that

THIA communicates in a manner participants found appropriately empathetic and easy to understand.

THIA also demonstrated a notable ability to overcome general skepticism toward AI. Participants entered the study with low baseline trust in AI-generated health information (M = 2.67 out of 5). Following interaction with THIA, trust rose to 4.00 – a gain of 1.33 points – which, in qualitative interviews, was attributed to NORC’s organizational backing and the platform’s alignment with participants’ caregiving experiences. Overall usability, as measured by the SUS, yielded a score of 80.42 out of 100, placing THIA in the Excellent category and the top 10% of platforms evaluated using this instrument. See Table 3 for complete performance results and trust development metrics.

**Table 3.** User Testing Performance Results (N = 6)

| Measure/Dimension | Pre | Post | Δ | Classification |
| --- | --- | --- | --- | --- |
| Technical Aspects | — | 4.72 | — | Excellent |
| Privacy & Data Security | — | 4.46 | — | Strong |
| User Experience | — | 4.37 | — | Strong |
| Usefulness & Relevance | — | 4.31 | — | Strong |
| Trust & Credibility | — | 4.21 | — | Strong |
| Trust: General AI | 2.67 | — | — | — |
| Trust: THIA-specific | — | 4.00 | +1.33 | Strong (+49.8%) |
| System Usability (SUS, 0-100) | — | 80.42 | — | Excellent (top 10%) |

*All post-interaction measures on 5-point scale. SUS industry benchmark = 68. Pre-interaction baseline measured general AI trust; post-interaction measured THIA-specific trust following hands-on use.*

Backend analytics provided additional insight into platform performance under real-world conditions. A total of 38 user questions were asked by the six participants over the course of their sessions with THIA. THIA achieved a 91.7% success rate on caregiver emotional support queries and 87.5% on mental health resources – its primary content areas for this initial deployment. A notable finding emerged regarding citation behavior. Citations were present in 92% of THIA’s responses, yet click-through rates were 0%; most participants did not notice them because of their side-panel placement. However, when citations were noticed, they were the strongest individual trust driver in the survey (M = 4.50 out of 5), a finding corroborated across quantitative survey data, qualitative interviews, and backend analytics.

## Discussion

The sequential design of this evaluation framework demonstrated how adversarial red-teaming and structured user testing surface distinct categories of platform risk that neither method could identify alone. Red-teaming, conducted first, identified and enabled the resolution of platform integrity issues before any member of the target population interacted with THIA. User testing, conducted on the improved platform, surfaced dimensions of platform readiness – trust formation, real-world use patterns, and emotional response – that adversarial testing was not designed to capture. Together, the two methods produced an evidence base sufficient to support refinement and deployment decisions, institutional review board assessment, items for ongoing monitoring, and processes for documentation and reporting.

The specific issues identified through this process illustrate this point. Red-teaming surfaced PII echoing in responses, system instability under high-input conditions, and inconsistent citation formatting – all remediated before end users interacted with the platform. User testing then identified a distinct category of risk: citations were present in 92% of responses and accurate, but their side-panel placement resulted in a 0% click-through rate because users simply didn’t notice them. That each evaluation phase surfaced what the other could not – and that remediation occurred between phases – is what distinguishes a sequential framework from standalone testing.

The framework presented in this paper demonstrates how a single structured evaluation can surface distinct concerns and address different questions across both platform integrity and user readiness. A platform may hold appropriate clinical boundaries under adversarial testing while still failing to build trust with real users – or may resonate strongly with users while harboring safety vulnerabilities that real-world interaction is unlikely to expose. The integration of both methods into a sequential pre-deployment protocol helps ensure that risks are properly identified and mitigated before deployment with end users. As such, it reflects the dual obligations of responsible AI deployment in public health: genuinely serve the intended population while minimizing risks. The methods, instruments, and scoring rubrics described in this report are documented to support adaptation by other public health organizations developing or deploying AI-powered health communication tools.

### Limitations

Several limitations of this study warrant acknowledgment. Sample sizes were small – five participants in red-teaming and six in user testing – reflecting the practical constraints of an internally-funded pre-deployment evaluation. Additionally, two-thirds of user-testing participants reported prior experience with AI tools, which may have contributed to higher baseline comfort than AI-naive users. While this proportion may reflect growing AI adoption among health information seekers, the findings may not generalize fully to caregivers with no prior exposure to AI. The evaluation framework described here has been used in a single deployment context with one population and one topic area; While the methods, instruments, and scoring rubrics shared here are designed for adaptation across platforms, topics, and populations to inform practice in the field, their applicability in other deployment contexts remains to be tested. Longitudinal evaluation across subsequent THIA deployments and larger and more diverse populations will be necessary to assess the framework’s broader utility and refine methods over time.

## Conclusion

The dual-method framework described here offers a practical, evidence-based model for responsible pre-deployment risk evaluation of AI tools in public health research. THIA’s strong performance across both evaluations – and the identification and resolution of priority issues throughout the process – demonstrates that structured pre-deployment evaluation is both feasible and consequential for public health AI tool deployment.

As generative AI transitions from novelty to infrastructure in public health research and practice, the field requires shared standards for what responsible pre-deployment risk evaluation entails. This paper proposes and documents one such standard – sequential, dual-method, grounded in applied practice, and oriented toward the populations that public health AI tools are designed to serve.

### Implications for Policy and Practice

This report offers three contributions for practitioners and the field:

- **A preliminary methods framework**: public health organizations deploying AI communication tools should adopt structured pre-deployment risk evaluation protocols that address both platform integrity and user readiness before engaging study participants or client populations. The dual-method framework described here offers one such model.
- **A reporting model**: standardized reporting enables comparison across tools and deployments over time, and supports the accountability that responsible AI use in public health requires. The documentation structure used here provides a template for organizing and communicating pre-deployment evaluation findings to funders, institutional review boards, and research partners.
- **A transparency standard**: funders and institutional review boards should treat documented, publicly reported pre-deployment evaluations as a condition of AI tool deployment in public health research and practice settings. By publishing this evaluation – including what was found, what failed, and what was fixed – this paper models the transparency that the field should expect and require. In the absence of field-wide mandates, early adopters who publish their evaluation findings establish the norm. This paper is one such contribution.

## Data Availability Statement

The datasets generated and/or analyzed during the current study are not publicly available due to participant confidentiality but may be available from the corresponding author upon reasonable request.

## Acknowledgment

Use of Artificial Intelligence: The authors used generative artificial intelligence tools to assist with manuscript outlining, editing, and language refinement. All AI-generated content was reviewed, verified, and revised by the authors, who take full responsibility for the accuracy, interpretation, and final content of the manuscript.

