## Supplemental Material 3 for "Toward Standards for Responsible AI in Health Communication: A Dual-Method Pre-Deployment Risk Evaluation Framework and Case Study"

**Section 1: Consent & Introduction (1 min)**
**Welcome Page:**
**Title:** THIA User Testing Study

Thank you for participating in this user testing study for THIA (Trusted Health Information Assistant), NORC's AI-powered platform designed to provide trustworthy health information for caregivers of older adults. This complete session (including interaction with THIA) will take approximately 30-40 minutes. Your participation is voluntary, and you may withdraw at any time. Your responses will be kept confidential and used only for research purposes. Upon completion, you will receive a $20 Amazon gift card.

**Consent Question:**

1. **Do you agree to participate in this study?**
    - Yes, I agree to participate [Must select to move forward]
    - No, I do not wish to participate [Skip to end]

---

**Section 2: Demographics (2 min)**

2. **Which of the following age ranges are you in?**
    - 18-24 years
    - 25–34 years
    - 35–44 years
    - 45–54 years
    - 55–64 years
    - 65+ years
3. **Which of the following best describes your current gender identity? (Select all that apply)**
    - Male
    - Female
    - Non-binary
    - Transgender
    - Prefer to self-describe: [text box]
    - Prefer not to answer
4. **What is your race? (Select all that apply)**
    - ❑ Asian or Asian American
    - ❑ Black or African American
    - ❑ Native American or Alaska Native
    - ❑ Native Hawaiian or Pacific Islander
    - ❑ White/Caucasian
    - ❑ Other: [text box]
    - ❑ Prefer not to answer
5. **Are you of Hispanic, Latino, or Spanish origin?**

- Yes
- No
- Prefer not to answer

6. **What is your highest level of education completed?**
    - High school or less
    - Some college
    - Associate's degree
    - Bachelor's degree
    - Graduate or professional degree

**Caregiving Background:**

7. **How long have you been a caregiver to an older adult?**
    - Less than 6 months
    - 6 months to 1 year
    - 1-3 years
    - 3-5 years
    - More than 5 years
8. **Who are you caring for?**
    - Parent
    - Spouse/Partner
    - Grandparent
    - Other family member
    - Friend/Neighbor
    - Other: [text box]
9. **On average, how many hours per week do you spend providing care?**
    - Less than 5 hours
    - 5-10 hours
    - 11-20 hours
    - 21-40 hours
    - More than 40 hours

**Technology Use:**

10. **How often do you use the internet?**
    - Multiple times per day
    - Once per day
    - A few times per week
    - Once per week or less
    - Rarely/Never
11. **Have you ever used AI tools (such as ChatGPT, Claude, Gemini, Alexa, Siri, or similar)?**
    - Yes
    - No [Skip to Q13]
12. **[If YES to Q11] Which AI tools have you used? (Select all that apply)**
    - ❑ ChatGPT
    - ❑ Google Bard/Gemini
    - ❑ Claude.ai

- ❑ Microsoft Copilot
- ❑ Alexa
- ❑ Siri
- ❑ Other: [text box]

---

**Section 3: Pre-Interaction Questions (1 min)**
**Health Information Seeking:**

13. **How often do you search for health information online?**
    - Daily
    - Several times per week
    - Once per week
    - A few times per month
    - Rarely
    - Never
14. **Where do you typically look for health information? (Select all that apply)**
    - ❑ Search engines (Google, Bing, etc.)
    - ❑ Health organization websites (CDC, NIH, Mayo Clinic, etc.)
    - ❑ Social media (Facebook, Instagram, TikTok, etc.)
    - ❑ AI chatbots (ChatGPT, etc.)
    - ❑ Healthcare provider
    - ❑ Friends or family
    - ❑ Other: [text box]
15. **How much do you trust AI-generated health information?**
    Scale: 1 (Not at all) to 5 (Completely)

16. **Have you ever used AI tools specifically to find caregiving information?**
    - Yes [Skip to Q17]
    - No [Skip to Section 4]
    - Not sure
17. **Provide an example of the type of caregiving information you search for when using AI tools?**
    [Open-ended question]

---

**Section 4: THIA Interaction Instructions (3-4 min)**

**Instruction Page:**
**"Now you'll interact with THIA (Trusted Health Information Assistant)"**
THIA is an AI-powered chatbot designed to provide trustworthy health information for caregivers of older adults. It draws from vetted health sources and provides citations for its responses.

**Your Task:**
Imagine you're feeling overwhelmed as a caregiver and need support. Please:

1. Click the link below to access THIA. You’ll be prompted to *log in with the username and password* I will provide you with.

2. Spend 15-20 minutes exploring the tool and having a natural conversation. Feel free to ask any questions about **caregiving challenges, mental health concerns, or resources you might need as a caregiver.** Examples of topics you might explore:
    - Managing caregiver stress or burnout
    - Finding mental health support
    - Coping with difficult emotions related to caregiving
    - Resources for specific caregiving situations
    - Self-care strategies
3. Pay attention to the information provided, the interface design, and how easy it is to use

There are no right or wrong questions—just interact as you naturally would if you needed caregiving support. When you're finished, click 'Next' to continue with the survey.

>> INSERTED THIA LINK HERE<<

---

**Section 5: Post-Interaction Feedback (4-5 min)**

**System Usability Scale (SUS)** *(5-point Likert Scale: 1=Strongly Disagree to 5=Strongly Agree)*
"Please rate your level of agreement with the following statements about THIA:"

18. I think that I would like to use THIA frequently
19. I found THIA unnecessarily complex
20. I thought THIA was easy to use
21. I think that I would need the support of a technical person to be able to use THIA
22. I found the various functions in THIA were well integrated
23. I thought there was too much inconsistency in THIA
24. I would imagine that most people would learn to use THIA very quickly
25. I found THIA very cumbersome to use
26. I felt very confident using THIA
27. I need to learn a lot of things before I could get going with THIA

**User Experience** *(5-point Likert Scale: Strongly Disagree to Strongly Agree)*
Please rate your level of agreement with the following statements:

28. The interface was clear and intuitive
29. I could easily find the information I was looking for
30. THIA responded to my questions in a timely manner
31. The conversation with THIA felt natural
32. I felt comfortable sharing my caregiving concerns with THIA

**Trust & Credibility** *(5-point Likert Scale: Strongly Disagree to Strongly Agree)*

33. I trust the information THIA provided
34. The source citations increased my confidence in the responses
35. THIA's responses seemed accurate and evidence-based
36. THIA seemed like a reliable source of health information

**Usefulness & Relevance** *(5-point Likert Scale: Strongly Disagree to Strongly Agree)*

37. The information provided was relevant to my needs as a caregiver

38. The conversations with THIA were helpful in expressing my caregiving experiences
39. THIA's tone was empathetic and appropriate
40. The information was presented in a way that was easy to understand
41. THIA provided actionable advice I could use
42. I would use THIA again for health or caregiving information

**Technical Aspects** *(5-point Likert Scale: Strongly Disagree to Strongly Agree)*

43. THIA understood my messages correctly most of the time
44. I rarely experienced technical issues while using THIA
45. The chat history was well-organized and easy to review

**Privacy** *(5-point Likert Scale: Strongly Disagree to Strongly Agree)*

46. I felt my personal information was secure when chatting with THIA
47. I trusted THIA's ability to handle sensitive information
48. I was comfortable with how my conversation data was being used
49. The privacy policy and data usage were clearly explained

**Comparative Questions:**

50. Compared to other sources you use for health information (Google, healthcare providers, etc.), how would you rate THIA?
    - Much worse
    - Somewhat worse
    - About the same
    - Somewhat better
    - Much better
51. How likely are you to recommend THIA to other caregivers?
    - 0 (Not at all likely) to 10 (Extremely likely)

**Open-Ended Questions:**

52. What did you like most about THIA?
    - [Large text box, 3-5 sentences]
53. What could be improved about THIA?
    - [Large text box, 3-5 sentences]
54. How did THIA help or hinder your caregiving journey?
    - [Large text box, 3-5 sentences]
55. Were there any topics or concerns you wished THIA could address better?
    - [Large text box, 3-5 sentences]
56. Is there anything else you'd like to share about your experience with THIA?
    - [Large text box, 3-5 sentences]

**Overall Assessment:**

57. Overall, how satisfied are you with THIA?
    - Very Dissatisfied
    - Dissatisfied
    - Neutral
    - Satisfied

- Very Satisfied

---

**Section 6: Incentive & Closing (1 min)**
**Thank You Page:**
Thank you for participating in the THIA user testing study! Your feedback is invaluable and will help us improve THIA to better serve caregivers like you.

**To receive your $20 Amazon gift card:**
Please provide your preferred email address and gift card preference below.

**Email address:** [text box]

You should receive your gift card within 5-7 business days.
If you have any questions or concerns, please contact.
Thank you again for your time and insights!
