## Supplemental Material 4 for "Toward Standards for Responsible AI in Health Communication: A Dual-Method Pre-Deployment Risk Evaluation Framework and Case Study"

**Estimated Duration:** 30 minutes
**Incentive:** $10 Amazon gift card

---

**Introduction (2 minutes)**
**Script:**Thank you for agreeing to this follow-up interview. As you know, I'm Neha Trivedi from NORC, and I'm working on the THIA user testing project.

This interview will take about 30 minutes. The purpose is to dive deeper into your experience using THIA and get your detailed feedback on what worked well and what could be improved. There are no right or wrong answers. I'm just interested in your honest thoughts and experiences.

Everything you share will be kept confidential and used only for research purposes to improve THIA. With your permission, I'd like to record this conversation so I can focus on our discussion rather than taking notes. The recording will only be used by our research team and will not be shared outside NORC.

Do you consent to this interview being recorded?

- Yes, consent obtained
- No, take notes only

**Section 1: Overall Experience Reflection (5 minutes)**

1. Let's start by thinking back to when you first opened THIA. Can you walk me through what you did first?
   - *Probe: Can you walk me through some of the questions or topics you explored with THIA?*
   - *Probe: What did you ask about? Why did you choose those topics?*
2. Can you tell me about your overall experience using THIA? What stands out most in your memory?
   - *Probe: What was your first impression when you started using it?*
3. Thinking back to when you interacted with THIA, what was your emotional response—how did it make you feel?
   - *Probe: Comfortable? Frustrated? Supported? Skeptical?*

---

**Section 2: Interaction Details & Content Quality (8 minutes)**

3. How satisfied were you with the answers THIA provided?
   - *Probe: Were the responses helpful? Accurate? Relevant to your needs?*
   - *Probe: Were there times when THIA didn't understand your question or gave an unhelpful response?*
   - *Probe: THIA provides source citations for its responses. Did you notice these? Did they affect your trust in the information?*
4. Did THIA provide information you didn't already know or hadn't considered?

- *Probe: Can you give an example?*

5. You mentioned in your survey that [reference specific survey response]. Can you tell me more about that?
    - *Note: Customize this based on their survey responses—especially open-ended comments*

---

**Section 3: Interface (7 minutes)**

6. Now let's talk about the visual design of THIA. What did you think about how THIA looked, things like the colors, logo, fonts, and overall visual style?
    - *Probe: Did anything stand out to you visually—either positively or negatively?*
    - *Probe: Was the interface visually appealing?*
    - *Probe: How did the citations appear? Were they easy to notice and read?*
3. Did you experience any technical issues during your interaction?
    - *Probe: Slow responses? Errors? Freezing? Unclear formatting?*
4. What did you think about the tone and style of THIA's responses?
    - *Probe: Did it feel conversational? Too formal? Empathetic? Robotic?*
5. If you could change one thing about the interface or how THIA works, what would it be?

---

**Section 4: Trust, Privacy & Safety (4 minutes)**

6. How much did you trust the information THIA provided?
    - *Probe: What made you trust it (or not trust it)?*
    - *Probe: How does it compare to other sources like Google, your doctor, or health websites?*
7. Did you have any concerns about privacy or how your data might be used?
    - *Probe: Would you feel comfortable sharing sensitive or personal health information with THIA?*
    - *Probe: What would help you feel safer?*

---

**Section 5: Value & Future Use (3 minutes)**

8. Can you imagine using THIA again in the future? In what situations?
    - *Probe: What would make you want to use it? What would prevent you?*

---

**Section 6: Suggestions for Improvement (3 minutes)**

9. If you were on the team developing THIA, what would you prioritize improving first?
10. Is there anything else about your experience with THIA that we haven't covered but you think is important for us to know?

---

**Closing (1 minute)**

Thank you so much for taking the time to share your detailed feedback. Your insights are incredibly valuable and will directly help us improve THIA to better serve caregivers.
As a reminder, you'll receive a $10 Amazon gift card within 5-7 business days to the email address you provided. If you think of anything else you'd like to share, please feel free to reach out to.

Do you have any questions for me?
